# A Data-driven Framework for Learning and Visualizing Characteristics of Thrombotic Event Phenotypes from Clinical Texts

**DOI:** 10.1101/2021.03.09.21253233

**Authors:** Anahita Davoudi, Audrey Yang, Sy Hwang, Danielle L. Mowery

## Abstract

Automatically identifying thrombotic phenotypes based on clinical data, particularly clinical texts, can be challenging. Although many investigators have developed targeted information extraction methods for identifying thrombotic phenotypes from radiology notes, these methods can be time consuming to train, require large amounts of training data, and may miss subtle textual clues predictive of a thrombotic phenotype from notes beyond the radiology note. We developed a generalizable, data-driven framework for learning, characterizing, and visualizing clinical concepts from both radiology and discharge summaries predictive of thrombotic phenotypes.

## Introduction

Generally-speaking, thrombotic events, *blood clots within the veins or arteries, obstructing blood flow through the body*, are a significant health threat to patients. In the United States, more than 795,000 individuals experience a stroke,^1^ 900,000 people experience venous thromboembolism,^2^ and 1.5 million people experience myocardial infarctions each year.^3^ Electronic health records afford an opportunity to study thrombotic phenotypes i.e., their disease burden, treatment efficacy, and health outcomes, among patients as they contain rich details about a patient’s clinical status including signs and symptoms, comorbidities, laboratory findings, procedures, and diagnoses.

### Natural Language Processing to Detect Thrombotic Phenotypes

Natural language processing (NLP) can accurately identify patients with thrombotic event phenotypes using patient’s clinical notes. Regular expression and rule-based approaches can extract and encode thrombotic events in an intuitive and explainable way. The pyConText algorithm, a regular expression and rules-based algorithm for identifying thrombotic event targets (e.g., pulmonary embolisms, deep vein thrombosis) and their contexts (existence, uncertainty, acuity), has demonstrated promising results.^4–6^ Specifically, pyConText detects pulmonary embolisms from CT pulmonary angiography reports with recalls (and precisions) of 98% (83%), 86% (96%), 94% (93%), and 60% (90%) for disease state (pulmonary emboli present or absent), quality state (diagnostic or not diagnostic), certainty state (uncertainty present or absent), and temporal state (acute or chronic), respectively.^4^ The pyConText knowledge base was further developed to detect deep vein thrombosis as well as stroke and its risk factors.^5–10^ Moreover, using the web-based query-building tool called Data Discovery and Query Builder (DDQB), Tien et al. have shown how such expressions and rule-logic for negation can be leveraged to classify thrombotic events 30-days following hip and knee surgeries.^11^ Their rule-based approach achieved high results: recall (97%) and specificity (99%) for deep vein thrombosis, recall (97%) and specificity (100%) for pulmonary embolism, and recall (100%) and specificity (99%) for myocardial infarction. Some NLP systems leverage standardized vocabularies. The Reveal NLP Engine, based on the MedLEE (Medical Language Extraction and Encoding Systems), extracts clinical terms using the Systematized Nomenclature of Medicine (SNOMED) terminology and applies inference rules to classify VTEs from radiology reports.^12^ Reveal NLP has demonstrated high sensitivity (83%) and specificity (97%) when processing 6373 radiology reports from 3,371 hospital encounters.

Advanced deep learning approaches have furthered thrombotic event phenotype detection. Intelligent (context-aware) Word Embedding (IWE) utilizes domain-specific semantic dictionary mappings to train a neural embedding to detect pulmonary embolisms documented within chest CT radiology reports.^13^ IWE performed comparably to pyConText on the UPMC dataset with an F1-score of 94% to 92% which was used originally to tailor the pyConText model. Johnson et al. developed an NLP pipeline using a semi-automated binary labeling for encoding radiology notes indicating patients with and without pulmonary embolism.^14^ Initially, a rule-based method has been used to scan the radiology reports for the existence of a set of pre-defined regular expressions related to the lack of PE evidence in the report. A pre-trained BERT model was then fine-tuned on the training subset of the data, which led to 99% accuracy in predicting correct labels. Ong et al. used a range of NLP-based techniques to detect the presence or absence of strokes including subtypes and characteristics of ischemic stroke, middle cerebral artery territory involvement, and stroke acuity in radiographic reports. They leverage a variety of word to vector transformation approaches, including bag-of-words, TF-IDF, and GloVe to train logistic regression, k-nearest neighbor, decision tree, random forest, and recurrent neural networks. Bag-of-words were observed to be more compatible with low variance classifiers such as logistic regression. In contrast, GloVe may perform better following deep learning approaches such as recurrent neural networks. Overall, the NLP pipeline achieved AUC-ROC in the range of 80% to 95% for the three different tasks.

Although many of these approaches demonstrate the benefits of using rule-based and supervised learning approaches to detect thrombotic phenotypes, these approaches can be time consuming to train, require large amounts of training data, and may miss subtle textual clues predictive of a thrombotic phenotype from notes beyond the radiology note. Furthermore, these approaches do not often address or characterize multiple types of thrombotic phenotypes at once. A data-driven framework for learning, characterizing, and visualizing clinical concepts associated with thrombotic phenotyped cohorts can be leveraged to overcome these limitations and uncover known and novel as well as common and distinct characteristics between each thrombotic phenotype. Our long-term goal is to study the disease burden and health outcomes among patients that experience thrombotic phenotypes as a result of COVID-19. Our short-term goal is to create a data-driven framework to 1) learn various text-based, clinical concepts predictive of thrombotic phenotypes across note types in an unbiased and automated fashion, 2) identify common and distinct clinical concepts from clinical notes for each thrombotic phenotype, 3) understand how well these concepts inform automatic document classification across thrombotic phenotypes, and 4) visualize semantic relationships among informative clinical concepts for clinical interpretation.

### Methods

This study was approved by the University of Pennsylvania Institute Review Board (#831895). We leveraged the Medical Information Mart for Intensive Care version 3 (MIMIC-III) database, a database consisting of de-identified, electronic health records for over 61,000 patients admitted to the Beth Israel Deaconess Medical Center in Massachusetts from June 2001 through October 2012.^15^ We queried the following MIMIC-III tables: patient, admission, ICD codes, and noteevent tables. We defined our thrombotic phenotype subgroups based on definitions from the Consortium for Clinical Characterization of COVID-19 by EHR (4CE) Acute Kidney Injury Working Group (see **Table 1**).^16^ Six subgroups of thrombotic phenotypes have been defined using the ICD-9 codes: *myocardial infarction, pulmonary embolism, stroke, arterial thrombosis, venous thromboembolism*, and *disseminated intravascular coagulation*.

**Table 1.**
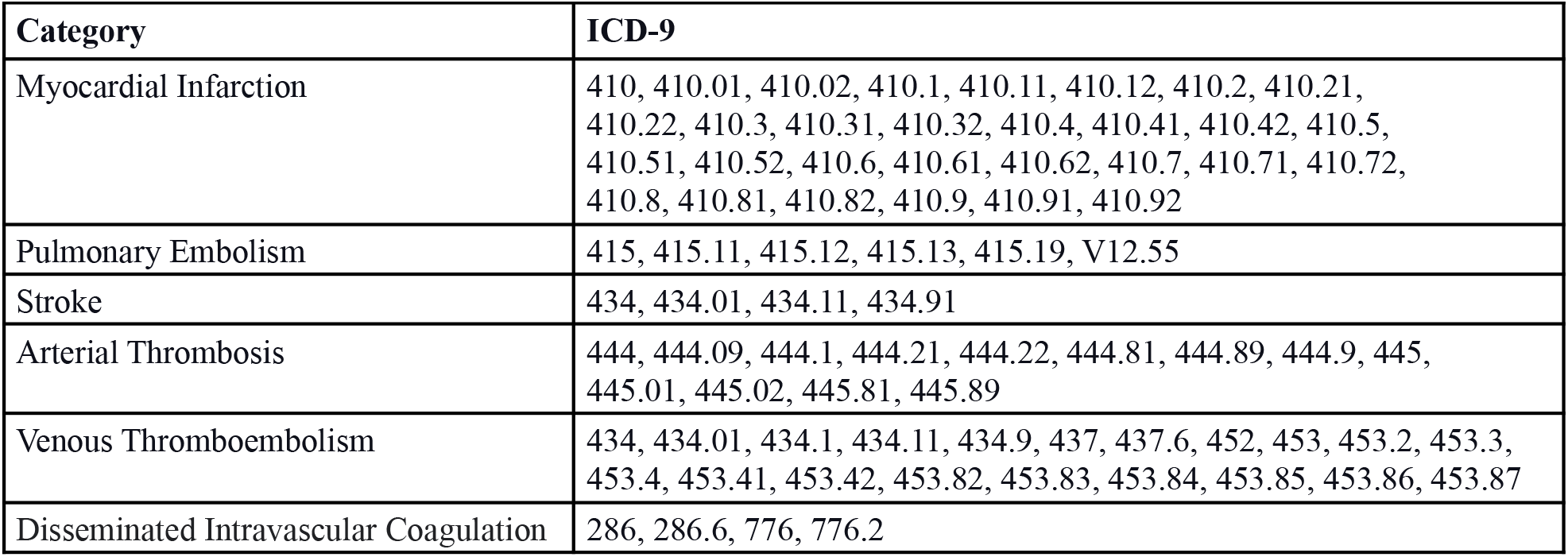
Thrombotic phenotype definitions.

### Applying a Text-driven Approach

We applied an NLP and machine learning-based method for learning distinct characteristics within each specific thrombotic phenotype using textual features from clinical notes. First, we leveraged discharge summaries and radiology notes in an effort to derive symptoms, signs, medications, procedures, diagnoses among other clinical concepts highly-associated with each thrombotic phenotype. Each patient may have one or more notes that are not associated with a thrombotic phenotype; therefore, we identified relevant notes containing terms associated with thrombotic phenotypes including: “thrombosis”, “thrombotic”, “thrombi”, “blood clot”, “blood clots”, “clot”, “clots”, “ischemia”, “ischemic”, “infarction”, “infarctions”, “infraction”, “infractions”, “embolism”, “embolisms”, “embolus”, “emboli”, “embolic”, “infarct”, “infract”, “occlusion”, “block”, “dissection”, “stroke”. For each patient, each of their individual notes were assigned the thrombotic event category associated with the ICD-9 billing code for that inpatient encounter. Next, we encoded clinical concepts identified using scispaCy’s Entity Linker.^17^ For each identified entity, we selected the top-ranked candidate among concepts mapped to standardized vocabularies including the Unified Medical Language System^18^, the Human Phenotype Ontology (HPO)^19^, and RxNorm. To identify the most informative clinical concepts across the full corpus, we applied the term frequency-inverse document frequency (TF-IDF), a measure that increases proportionally as the number of times a concept appears in a document relative to the number of documents that contain the concept. We report the top 20-ranked, positive-associated concepts for each thrombotic phenotype. To identify common and distinct clinical concepts learned between note types, we compared the clinical concepts learned between note types using jaccard similarity.

### Classifying Notes by Thrombotic Phenotypes

From each TF-IDF corpora, we selected the 2,000 most informative concepts to train and test a binary logistic regression model for classifying notes by thrombotic phenotype. The model was trained using 80% of notes and tested using 20% of notes. We applied 5-fold cross validation and L2 regularization to reduce the risk of overfitting. We report the feature importance of the top 20-ranked positive coefficients for each thrombotic phenotype. We also report F1-score, recall, and precision for both training and testing sets.

### Visualizing Clinical Concepts Associated with Thrombotic Phenotypes

We aimed to better understand the relationships between learned clinical concepts by applying an unsupervised clustering and visualization technique to explore all positive coefficients for each thrombotic phenotype. As a knowledge base of clinical concepts and their relationships, we leveraged cui2vec, a combined embedding resource from three medical data sources: insurance claims database of 60 million members, a collection of 20 million clinical notes, and 1.7 million full text biomedical journal articles resulting in 108,477 medical concepts.^20^ To visualize and observe semantic clusters among the learned clinical concepts for each thrombotic phenotype, we leveraged UMAP, a dimension reduction algorithm commonly used to reduce data representations into 2-dimensional space.^21^ As a proof-of-concept, we report relevant themes observed among tightly grouped concepts for each thrombotic phenotype.

## Results

In this pilot study, we aimed to develop a data-driven framework to identify, characterize, and visualize clinical concepts associated with six thrombotic phenotypes.

### Applying a Text-driven Approach

In **Table 2**, the most frequent thrombotic phenotypes observed in our cohort include myocardial infarction (n=4714 patients), venous thromboembolism (n=1798 patients), and pulmonary embolism (n=1131 patients). Among thrombotic phenotypes, the most documents and corresponding CUIs were observed for myocardial infarction (n=10422 documents; n=27971 CUIs), venous thromboembolism (n=9666 documents; n=26326 CUIs), and pulmonary embolism (n=5068 documents; n=22442 CUIs). Thrombotic phenotypes with the most frequent positive coefficients within the logistic regression model were disseminated intravascular coagulation (n=708 CUIs), venous thromboembolism (n=700 CUIs), and arterial thrombosis (n=651 CUIs).

**Table 2.** Characteristics of patients, documents, and CUI counts during each filtering stage.

| Thrombotic phenotype | Patients | Documents | Total CUIs | Total CUIs with positive coefficients |
| --- | --- | --- | --- | --- |
| Myocardial Infarction | 4714 | 10422 | 27971 | 695 |
| Pulmonary Embolism | 1131 | 5068 | 22442 | 666 |
| Stroke | 687 | 4781 | 17570 | 545 |
| Arterial Thrombosis | 407 | 1761 | 14696 | 651 |
| Venous Thromboembolism | 1798 | 9666 | 26326 | 700 |
| Disseminated Intravascular Coagulation | 381 | 1498 | 14947 | 708 |

In **Figures 2a** and **2b**, the highest-ranked positive coefficients by *description* among thrombotic phenotypes across note types include: myocardial infarction (*heart attack, electrocardiogram: myocardial infarction finding, abnormal cardiac catheterization*), pulmonary embolism (*blood clot in artery of lung, pulmonary embolism*), stroke (*stroke, cerebrovascular accident*), arterial thrombosis (*endocarditis, ischemia, lower extremity, surgical incision*), venous thromboembolism (*stroke, cerebrovascular accident, blood clot in portal vein*), and disseminated intravascular coagulation (*slc25a10 gene, ascites, infection in the blood stream, discharge diagnosis*). Importance among the top 20-ranked, positive coefficients were higher among features within the discharge summaries alone compared to radiology + discharge summaries and radiology.

**Figure 2a.**
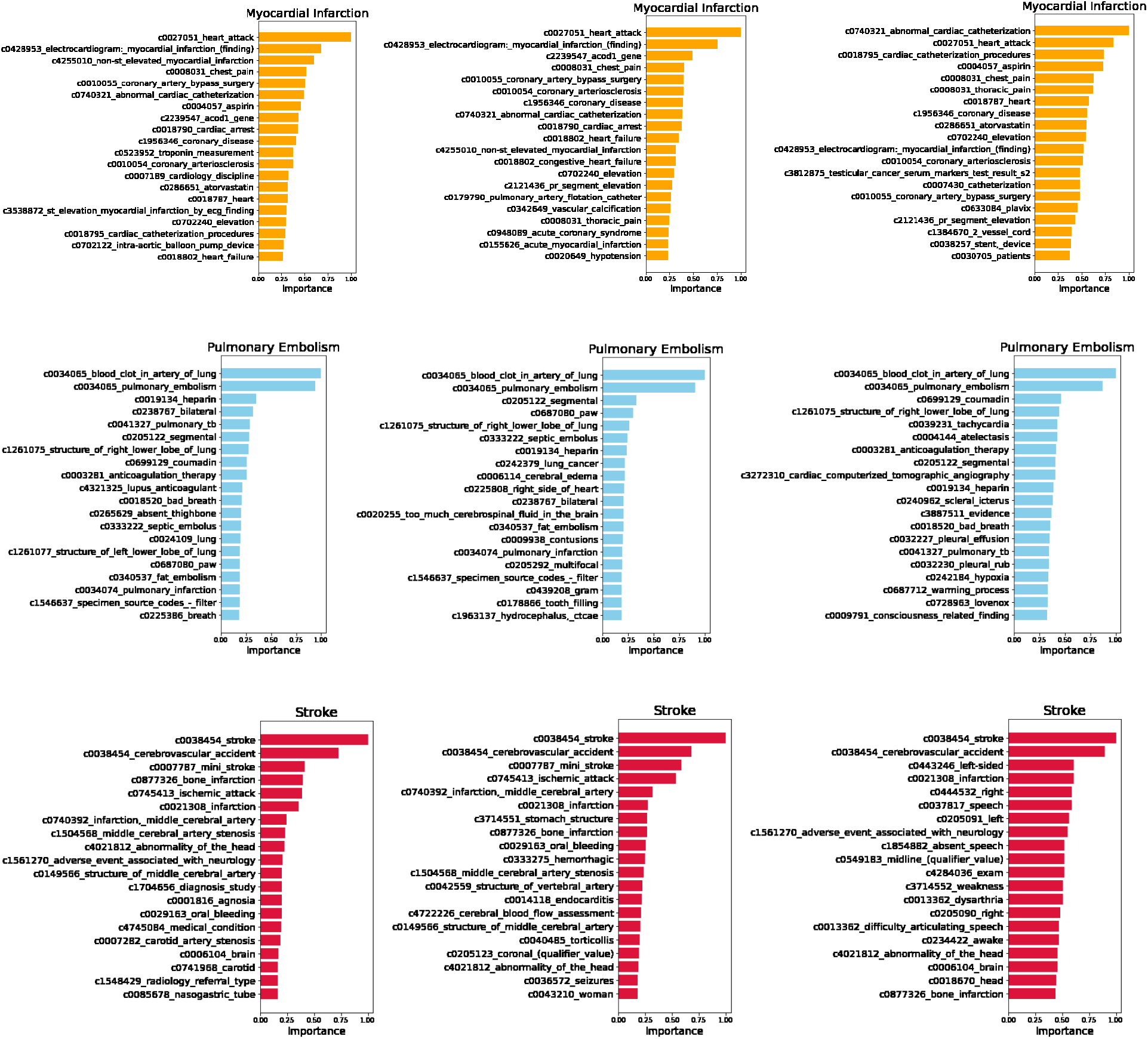
Feature importance of the top 20-ranked, positive features using logistic regression for each thrombotic phenotype. From left to right: *radiology + discharge summary, radiology*, and *discharge summary*.

**Figure 2b.**
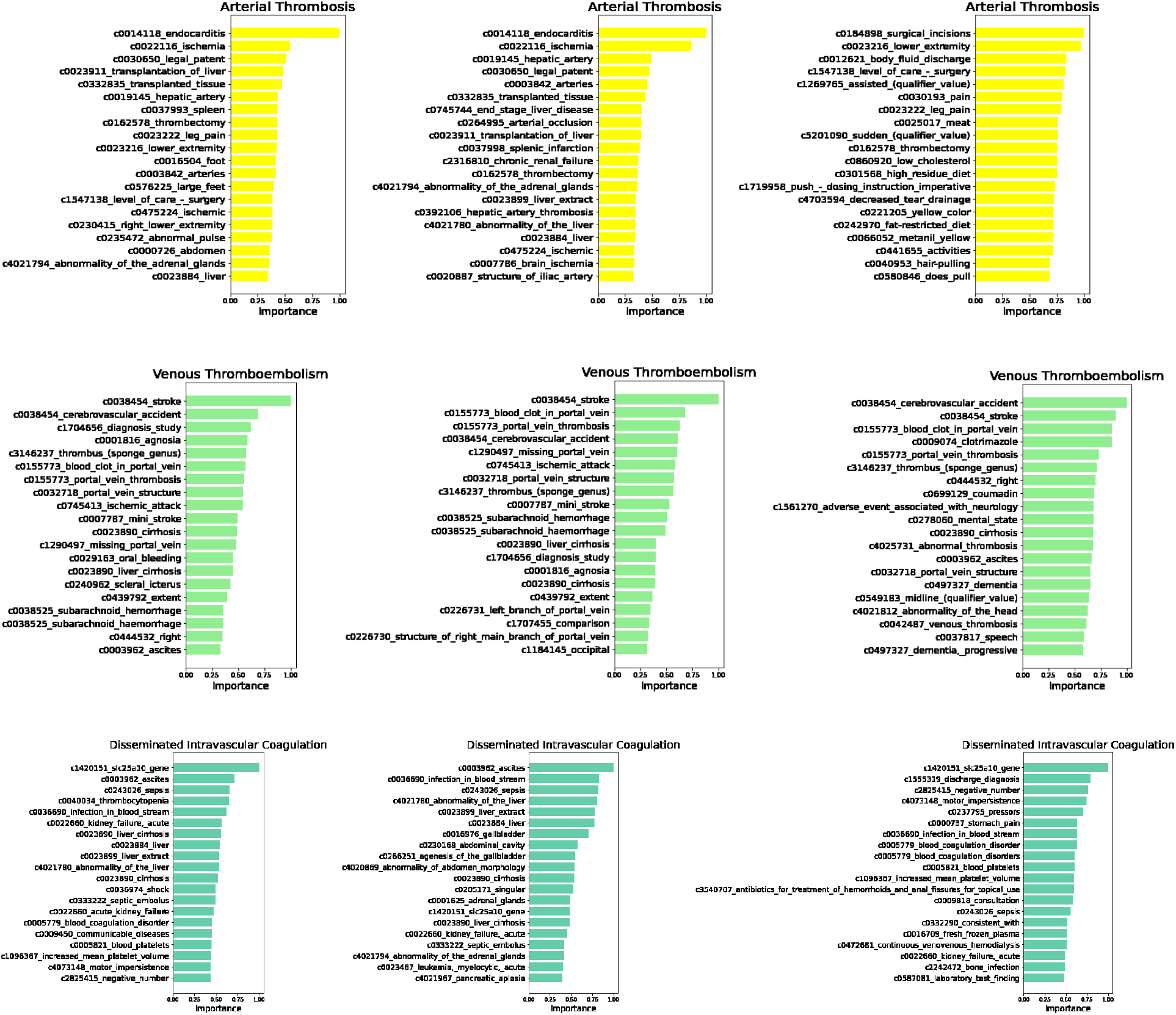
Feature importance of the top 20-ranked, positive coefficients using logistic regression for each thrombotic phenotype. From left to right: *radiology + discharge summary, radiology*, and *discharge summary*.

In **Figure 3**, we compared the clinical concepts learned across note types. We observed a range of common positive-associated clinical concepts from 0.446 (arterial thrombosis) to 0.505 (disseminated intravascular coagulation) between radiology + discharge summary and discharge summary; from 0.293 (myocardial infarction) to 0.335 (venous thromboembolism) between radiology + discharge summary and radiology; from 0.141 (myocardial infarction) to 0.206 (disseminated intravascular coagulation) between discharge summaries and radiology.

**Figure 3.**
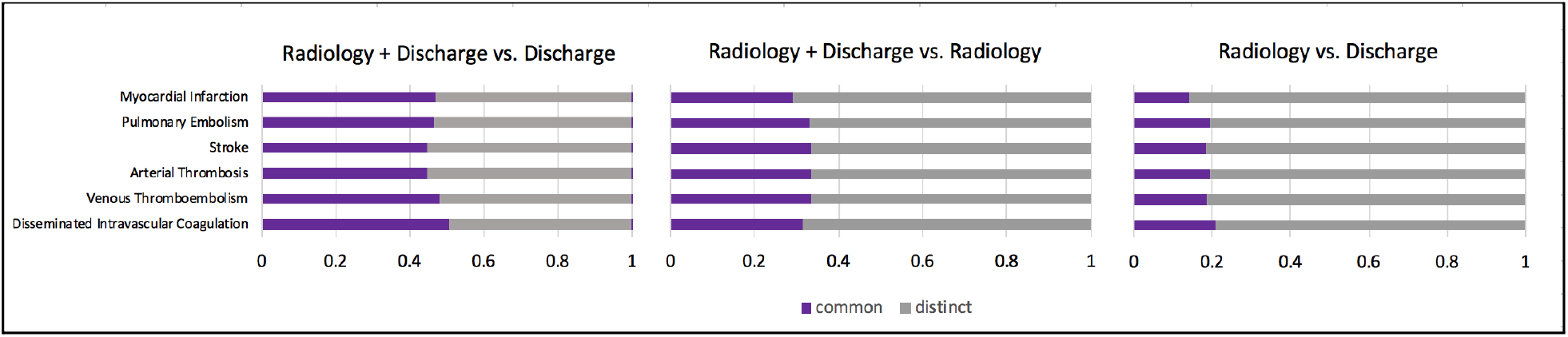
Jaccard similarity of clinical concepts between notes.

### Classifying Notes by Thrombotic Phenotypes

In **Table 3**, across thrombotic phenotypes, we observe slight reductions in performance, from high to moderate, on the testing set compared to the training set. On the testing set, we observe moderate to high F-score across thrombotic phenotypes and note types ranging from 0.63 to 0.82. The most sensitive models learned for myocardial infarction, pulmonary embolism, and venous thromboembolism were derived from discharge summaries; in contrast to, stroke, arterial thrombosis, and disseminated intravascular coagulation which were derived from radiology + discharge summaries. The most precise models learned for pulmonary embolism derived from radiology + discharge summaries; whereas, the most precise models for all other thrombotic phenotypes were derived from discharge summaries only.

**Table 3.** Logistic classification using TF-IDF. **Bold**=highest metric among note arms.

| Thrombotic phenotype | Training (n=80%) |  |  | Testing (n=20%) |  |  |
| --- | --- | --- | --- | --- | --- | --- |
| *Radiology + Discharge Summary* | *F-score* | *Recall* | *Precision* | *F-score* | *Recall* | *Precision* |
| Myocardial Infarction | 0.80 | 0.75 | 0.87 | 0.78 | 0.74 | 0.84 |
| Pulmonary Embolism | 0.80 | 0.80 | **0.81** | **0.79** | 0.76 | **0.79** |
| Stroke | 0.75 | 0.74 | 0.77 | 0.74 | **0.73** | 0.75 |
| Arterial Thrombosis | 0.75 | **0.78** | 0.72 | **0.73** | **0.77** | 0.69 |
| Venous Thromboembolism | 0.70 | 0.76 | 0.65 | 0.67 | 0.73 | 0.62 |
| Disseminated Intravascular Coagulation | 0.81 | 0.86 | 0.76 | **0.77** | **0.81** | 0.73 |
| *Radiology* | *F-score* | *Recall* | *Precision* | *F-score* | *Recall* | *Precision* |
| Myocardial Infarction | 0.76 | 0.74 | 0.78 | 0.70 | 0.67 | 0.73 |
| Pulmonary Embolism | 0.81 | 0.80 | **0.81** | 0.76 | 0.76 | 0.76 |
| Stroke | 0.75 | **0.75** | 0.75 | 0.72 | 0.71 | 0.73 |
| Arterial Thrombosis | 0.76 | 0.74 | 0.77 | 0.72 | 0.72 | 0.71 |
| Venous Thromboembolism | 0.65 | 0.66 | 0.65 | 0.63 | 0.65 | 0.62 |
| Disseminated Intravascular Coagulation | 0.70 | 0.67 | 0.75 | 0.66 | 0.62 | 0.69 |
| *Discharge Summary* | *F-score* | *Recall* | *Precision* | *F-score* | *Recall* | *Precision* |
| Myocardial Infarction | **0.83** | **0.79** | **0.88** | **0.82** | **0.77** | **0.87** |
| Pulmonary Embolism | **0.83** | **0.88** | 0.79 | **0.79** | **0.85** | 0.74 |
| Stroke | **0.81** | **0.75** | **0.87** | **0.78** | 0.71 | **0.86** |
| Arterial Thrombosis | **0.78** | 0.74 | **0.82** | **0.73** | 0.68 | **0.80** |
| Venous Thromboembolism | **0.81** | **0.87** | **0.76** | **0.75** | **0.81** | **0.71** |
| Disseminated Intravascular Coagulation | **0.86** | **0.88** | **0.83** | 0.76 | 0.76 | **0.76** |

### Visualizing Clinical Concepts Associated with Thrombotic Phenotypes

In **Figure 4**, among thrombotic phenotypes, using UMAP on cui2vec vectors demonstrates distinct patterns representing the preserved semantic relationships among concepts learned from clinical data sources.

**Figure 4.**
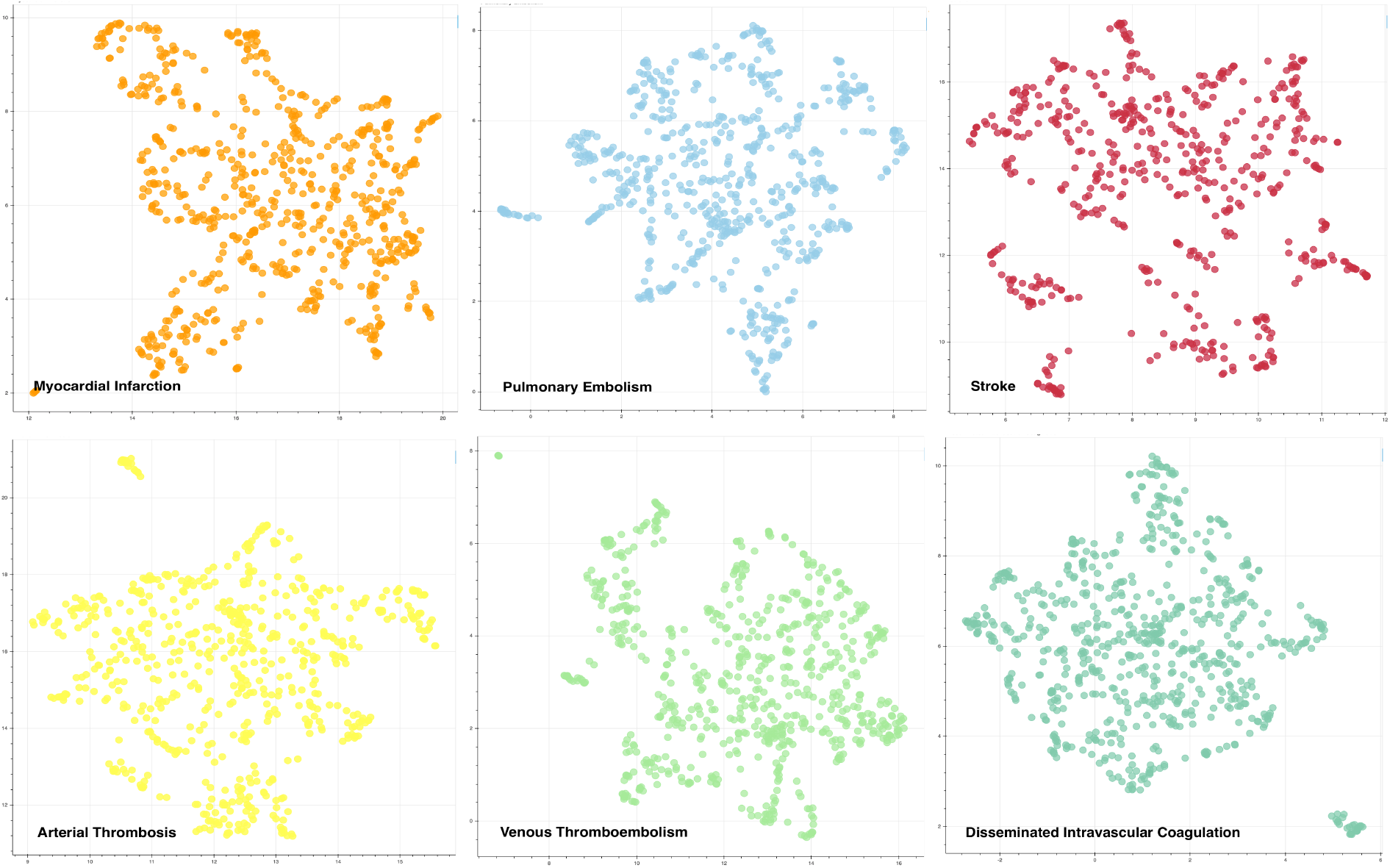
Visualization of clinical concepts with positive coefficients using UMAP on cui2vec vectors.

In **Figure 5**, within the UMAP visualizations for each thrombotic phenotype, we observed several interesting types of clinically-meaningful, semantic relationships including prophylaxis (myocardial infarction), comorbidities (pulmonary embolism), affected anatomy (stroke), care coordination (arterial thrombosis), synonyms (venous thromboembolism), biomarkers/treatments (disseminated intravascular coagulation).

**Figure 5.**
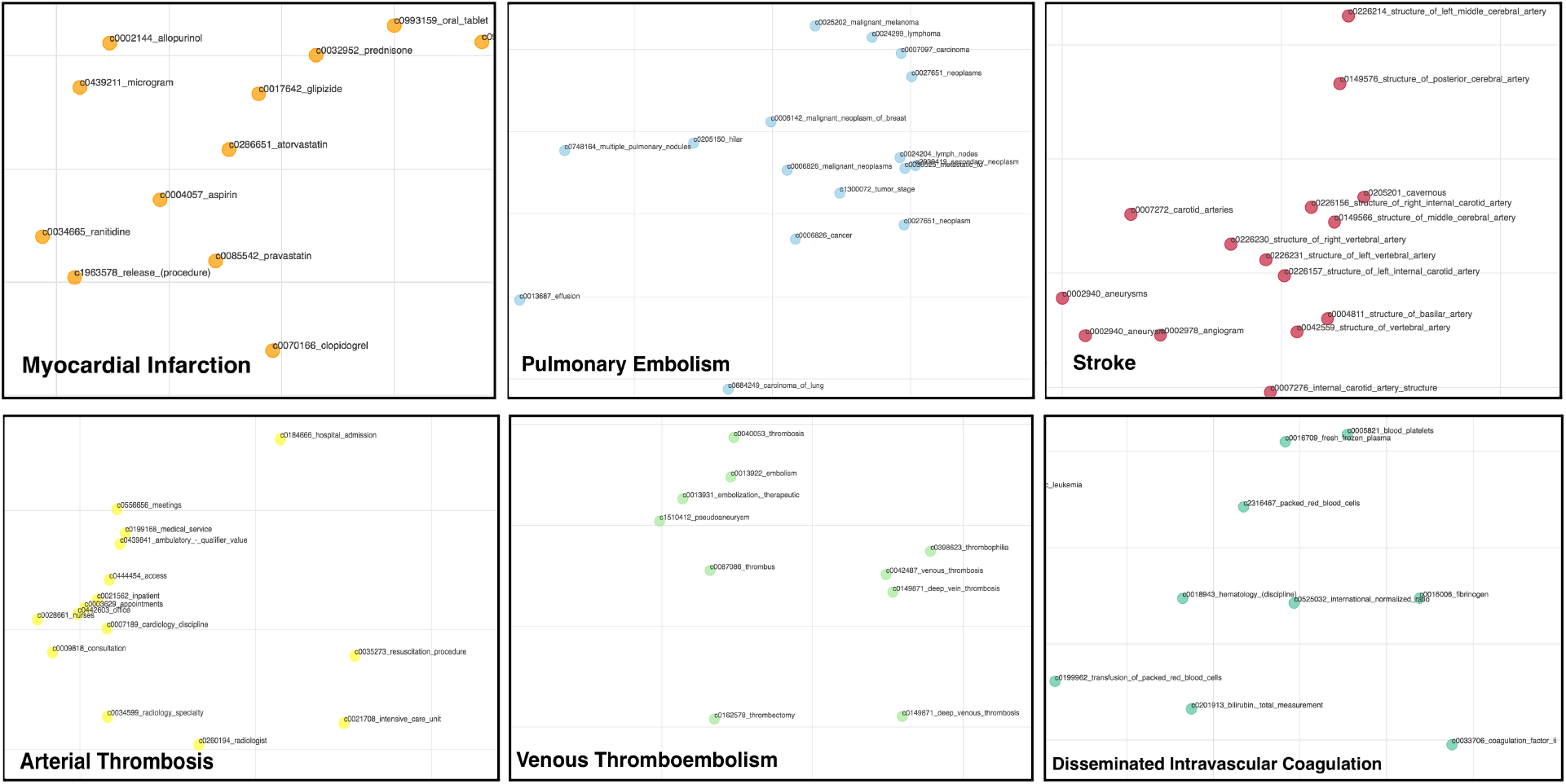
Semantic relationships observed by thrombotic phenotype.

## Discussion

In this pilot study, we developed and applied a data-driven framework to identify, characterize, and visualize clinical concepts associated with six thrombotic phenotypes.

### Applying a Text-driven Approach

We developed this data-driven approach using standard vocabularies and ontologies to glean informative clinical concepts from relevant note types - discharge summaries and radiology notes - for classifying documents according to thrombotic phenotypes. Among the top 20-ranked positive coefficients identified for specific thrombotic phenotypes of stroke, pulmonary embolism and myocardial infarction, both lay and medical synonyms of diagnoses were common, e.g., heart attack *is a* myocardial infarction. In the case of disseminated intravascular coagulation, we observed more heterogeneous, clinical concepts among the top-ranked. For example, pathogenic mechanisms (*infection in bloodstream also known as sepsis*) was highly predictive which is not surprising given that disseminated intravascular coagulation is a known complication of sepsis in about 35% of severe cases.^22^ In the case of arterial thrombosis, ischemia and endocarditis are also intuitive findings because endocarditis can cause vegetations of blood and bacteria to form in the blood vessels of your heart forming blood clots that may travel and cause ischemia within the arteries.^23^ Ascites fluid reinfusion in severe cirrhosis has frequently been associated with disseminated intravascular coagulation.^24^ Another intuitive finding by our method is that our model for arterial thrombosis identified affected arteries (*hepatic artery, arterial occlusion*); in contrast, our model for venous thromboembolism identified affected veins (*portal vein thrombosis, left branch of portal vein*). This finding demonstrates that our method can identify key vascular differences between these thrombotic phenotypes.

Across thrombotic phenotypes, the top 20-ranked, positive coefficients depicted from the discharge summary alone consistently appeared to have higher importance scores than those enumerated within the radiology + discharge summary or radiology note alone. Furthermore, when comparing the clinical concepts learned across note types, we observed higher jaccard similarity measures of positive-ranked clinical concepts between the discharge summary and radiology + discharge summary compared to radiology and radiology + discharge summary. This finding empirically demonstrates a higher proportion of clinical concepts were derived from the discharge summaries than the radiology note in the combined note model. Although the radiology report is often necessary for making a clinical diagnosis and mined for identifying affirmed thrombotic phenotypes, we learned that there are significant clinical indicators derived from the discharge summary which are important for detecting a thrombotic phenotype.

### Classifying Notes by Thrombotic Phenotypes

In terms of document classification of thrombotic phenotypes, we observed only slight reductions in performance on the testing set compared to the training set, suggesting that the cross-validation and L2 regularization did improve generalizability. On the testing set, we observe moderate to high F-score across thrombotic phenotypes demonstrating reasonable classification performance; however, additional features could improve both recall and precision. The variability of performance in terms of recall and precision across note types suggests that one model and note type might not be suitable across thrombotic phenotypes. We acknowledge that our approach could be improved by adding non-textual features indicative of a thrombotic phenotype i.e., laboratory data, hospital billing codes, and assessment scales. Furthermore, our approach does not consider the context of the concept (linguistic modifiers of negation, severity, temporality, and experiencer) which may improve precision of classifications.^9,25^

### Visualizing Clinical Concepts Associated with Thrombotic Phenotypes

We leveraged state-of-the-art word embedding resources and unsupervised dimension reduction techniques to encourage research teams (informaticists, clinicians, epidemiologists among others) to explore clinically meaningful, semantic relationships within and across thrombotic phenotypes.

### Limitations and Future Work

Our study has several notable limitations. First, encoding text to vocabularies is not perfect. Some features that were positively associated with a thrombotic phenotype are clearly errors, e.g., *slc25a10 gene* was associated with disseminated intravascular coagulation (DIC), but was likely a mapping omission to dicarboxylate ion carrier (DIC). To address this issue, we will apply more aggressive filtering techniques and acronym/abbreviation support.^26^ Second, our study aims to learn informative concepts common and distinct to thrombotic phenotypes across reports to train a document-level thrombotic phenotype classifier given that ICD coding can be imprecise. In the future, we will roll up classification to the patient encounter-level by training our model using physician-validated, thrombotic phenotypes applied to our COVID-19 patient cases.

### Conclusion

We defined a text-based, data-driven framework to learn, characterise, and visualize thrombotic phenotypes using clinical texts. This generalizable framework could prove beneficial for investigators interested in leveraging clinical notes to train a phenotype classifier, but not sure which features to include and which notes to generate their models.

## Data Availability

The cui embeddings can be found at: https://figshare.com/s/00d69861786cd0156d81. UMAP visualizations and CUIs with positive coefficients can be found at: https://github.com/semantica-NLP/Data_Driven_Thrombotic_Events.

https://figshare.com/s/00d69861786cd0156d81

## Acknowledgements

This work was funded by Dr. Mowery’s start-up funds through the University of Pennsylvania. We extend our gratitude to the open-source community for making their resources available. The cui embeddings can be found at: https://figshare.com/s/00d69861786cd0156d81. UMAP visualizations and CUIs with positive coefficients can be found at: https://github.com/semantica-NLP/Data_Driven_Thrombotic_Events.

